# Associations between county-level suicidality and cyberbullying in rural New York State youth are significantly mediated by sadness and self-harming: YRBS 2016-2023

**DOI:** 10.1101/2025.08.27.25334601

**Authors:** Laura E. Jones, Riya Nadkarni, Nicole Krupa

**Affiliations:** Center for Biostatistics, Bassett Research Institute, Bassett Medical Center One Atwell Road, Cooperstown, New York, United States; Columbia Mailman School of Public Health, Columbia University 722 W 168th St, New York, NY, United States

**Keywords:** YRBS, rural adolescent, suicidality, bullying, cyberbullying, sex, sexual orientation, sexual minority

## Abstract

**Background:** The ubiquity and popularity of social media among youth has led to skyrocketing increases in cyberbullying rates among American teens. National studies show cyberbullying victimization is experienced differently by sex, race, and sexual orientation. While rural youth experience bullying at higher rates than urban teens, few studies have focused on the rural experience.

**Aim:** A primary objective of this study is to assess associations between bullying modalities and suicidality among respondents to the Youth Risk Behavior Surveillance System Survey (YRBS) in a rural county in New York state and assess for effect modification by sex and sexual orientation. Secondly, we assess sadness and self-harming as mediators in relationships between cyberbullying or conventional bullying and suicidality.

**Methods:** We analyzed cross-sectional data from county-level YRBS, years 2016, 2018, 2021 and 2023, performing multivariable logistic regressions to assess adjusted relationships between cyberbullying, conventional school bullying and suicidality. Stratified models were used to test for effect modification by sex and sexual orientation and mediation analysis was performed to assess mediation of the relationships between suicidality and bullying modalities by self-harming or sadness.

**Results:** Bullied rural youth were significantly more likely to consider suicide, with larger associations for cyberbullying in girls, and conventional bullying in boys. Cyberbullying and conventional bullying were significantly associated with suicidality, with 55-63% mediation by self-harming, and 51-56% mediation by persistent sadness. Indirect effects were significantly moderated by sexual orientation, but not by sex.

**Conclusions:** Self-harming and sadness are significant mediators for suicidality in the presence of bullying modalities. Results suggest need for educational programs focused on bullying and responsible internet use alongside tailored interventions based on sex, sexual orientation, and mental health history.

## Introduction

As of 2022, a Pew survey showed nearly half of teens (ages 13 to 17) have experienced at least one form of cyberbullying behavior, with 28% experiencing multiple forms either online or via cellphone. Experiences of victimization differed by age, sex, and race of the subject (1), paralleling usage of online platforms that also varies by demographic (2). Cyberbullying involves use of technology to harass, humiliate, embarrass, or target a peer, and can take the form of online threats, false or rude texts or tweets, stalking, harassment, or sharing of personal information intended to harm or humiliate (3, 4). Unlike conventional bullying, cyberbullying allows perpetrators to remain anonymous and target victims regardless of location, making abuse harder to address (5). Consequences can be severe, as harmful content is visible to a wide audience who contribute to bullying by sharing or reposting it. Moreover, content can remain online indefinitely. Effects of cyberbullying on youth can be more severe than those resulting from traditional peer violence, as victims often lack control over the situation, leading to significant psychological harm despite absence of direct physical contact.

Low-income and rural youth are in general more susceptible to bullying, with students in rural United States experiencing bullying victimization at a higher rate (27.7%) than students in urban areas (22.4%)(6). Recent national statistics in the United states indicate that bullied rural youth are likelier to report cybervictimization (23.8%) than their urban counterparts (19%) (6). Urban-rural differences in school bullying and ‘poly-bullying’ (multiple forms of bullying, including direct and cyberbullying) victimization were found to be much higher in rural districts (20.3% for bullying; 7.7% for poly-bullying) than in urban areas (16.5% and 5.8%, respectively) in a 2024 study in Jiangsu Province, China (7).

Moreover, teen suicide has long been a greater problem in rural than in urban areas of the United States, with rates among both males and females nearly double those of urban youth (8). While suicides among American high school aged youth increased by 10% per year between 2014 and 2018, increases were higher among rural youth, with incidence among youth aged 10 to 19 years rising by 15% annually in rural areas (9) over the period from 2010 to 2018. Highly rural areas have fewer mental health facilities and suicide prevention services for young people than do urban areas. Only 3% of rural areas have mental health facilities providing specialized suicide prevention services for youths, in contrast to 8% of metropolitan areas (9).

Rural youth also have higher self-harm related outpatient and emergency medical visit rates compared to urban counterparts (10). Cyberbullying is associated with heightened risks of self-harm behaviors, including self-cutting or self-burning, among both victims and perpetrators (11). A recent (2020) study showed significant correlations between self-harming behaviors and increased risk of suicidal ideation among youth, and suggested that individuals with a prior history of self-injury were more likely to engage in suicidal acts (12). Depression (persistent sadness) and anxiety were found to significantly mediate associations between bullying and self-harm or suicidality in both sexes (13), with higher rates of depression and anxiety in girls.

Prevalences of suicidality (34.4% vs 21.6%) and self-harm (32.8% vs 22.3%) were higher in cyberbullying victims than those exposed to conventional school bullying (14) and youth subject to cyberbullying were more likely to self-harm and attempt suicide than those experiencing conventional direct or relational bullying at school (15).

Rates of bullying – and its effects – differ by sex, sexual orientation, and type of bullying experienced (16). A study using national YRBS data (2017) showed that cyberbullying and bullying victimization rates were higher among sexual minorities, with bisexual youth more vulnerable to bullying modalities than gay/lesbian students (17). Sexual minorities and non-conforming youth are at very high risk of cyberbullying relative to their heterosexual peers, with percentage of cyberbullying among LGBTQ youth ranging from 10.5% to 71.3% across 27 studies in a scoping review (18). Both male and female sexual minorities reported higher levels of cyberbullying than heterosexual counterparts, with female sexual minorities reporting higher rates of cyberbullying than male counterparts (19). Sexual minority youth who experience bullying and cyberbullying are also more likely to engage in substance abuse, as well as depressive, self-harming, or suicidal behavior (20).

Recent studies of national YRBS data from 2019 (N=13,605) showed significant associations between cyberbullying, conventional bullying, and both depression and suicidality, with stronger associations for students who experienced cyberbullying and strongest associations among students who reported both forms of bullying (21, 22). However, these recent studies focus on a single year and feature national YRBS records. Our present analysis of YRBS data from a long-term repository collected in Otsego County, Central New York, therefore offers insight into the relationship between bullying and adverse mental health indicators in under-studied rural youth.

The aims of this study are twofold: we first determine associations between suicidality and bullying modalities among rural youth respondents in the Otsego County, New York, with special attention to differences by sex and sexual orientation. Secondly, we examine the potential roles of sadness and self-harming as mediators in the relationships between cyberbullying or conventional bullying and suicidality.

## Methods and Materials

### Study Population

We analyzed cross-sectional data (N=4,846) from local Youth Risk Behavior Surveillance System (YRBS) surveys administered by Bassett Research Institute of Bassett Healthcare Network, Cooperstown, New York and focusing on youth from grades 9-12 in Otsego County for the years 2016, 2018, 2021 and 2023. YRBS monitors 6 categories of health behaviors, including behaviors contributing to unintentional injuries and violence: tobacco use; alcohol and other illicit drug use; sexual behavior, diet; and physical activity among high-school students in the United States. National and state level YRBS are conducted biennially in the form of self-administered questionnaires. Key variables for our study include questions addressing bullying and cyberbullying incidents, history of self-harming, and questions that document suicidal thoughts and plans.

### Exposures

Cyberbullying was assessed by the following question: “During the past 12 months, have you ever been electronically bullied?” and conventional school bullying was assessed with the question: “During the past 12 months, have you ever been bullied on school property?” Responses to school bullying were dichotomized to “yes” and “no” for any bullying in the prior 12-month period.

### Outcome

The main outcome variable is suicidality or suicidal ideation, a binary categorical variable where “yes” comprises any affirmative answer to either of the two following questions: “During the past 12 months, did you ever seriously consider attempting suicide?” and “During the past 12 months, did you plan about how you would attempt suicide?” Answers coded “no” in our binary variable thus require a negative response to both questions. YRBS metrics associated with suicidal ideation were found to have high convergent and discriminant validity (23).

### Mediators

Self-harming was assessed by the following question: “During the past 12 months, how many times did you do something to purposely hurt yourself without wanting to die, such as cutting or burning yourself on purpose?” Responses were dichotomized to binary 0/1 for any self-harming in the 12-month period. Sadness was assessed by the binary question, “During the past 12 months, did you ever feel so sad or hopeless almost daily for two weeks or more that you stopped usual activities?”

### Covariates

Covariates and potential confounders included grade (9 – 12 and ungraded), sex (male, female), sexual orientation (available since 2015: heterosexual, gay, bisexual, and not sure/questioning); and race-ethnicity (Hispanic, Native American, Black, Asian, White non-Hispanic and undeclared). A screen-time (hours per week) variable is available but is a potential mediator (Sobel test for mediation p = 0.003 with screen-time mediating cyberbullying; p = 0.04 with screen-time mediating bullying) (24) and thus was not included as a confounder.

## Statistical analysis

### Missing Data and Imputation

Missing values did not exceed 7.6% on any given covariate, but missing units resulted in a 38% loss to the size of our dataset, reducing it to 3005 samples from 4846 samples. We thus imputed our dataset using a fully conditional chained equations scheme employing predictive mean matching (PMM), and created 20 imputed datasets (25, 26) using the mice package in the R programming language (26). All analysis was performed on 20 imputed datasets, and the results pooled via Rubin’s Rules (27). Missing data are summarized in Table S1.

### Bivariate analysis

As a first step in assessing potential mediation, we conducted bivariate logistic regression analyses and used chi-square (χ²) tests to examine unadjusted associations between suicidality and exposures including self-harming, sadness, cyberbullying and traditional school bullying. Associations between demographic covariates including sex, race, and sexual orientation, and suicidality, self-harming, and bullying modalities also were assessed via likelihood ratio and chi-square (χ²) tests. All analysis was pooled over 20 imputed datasets.

Pooling of Chi-square tests results in a D_2_ statistic that is approximately F-distributed (28, 29) and see also (30) for a comparison of pooling methods.

### Multivariable analysis

We performed multivariable logistic regressions to assess relationships between exposure to bullying modalities and suicidality, testing for effect modification by sex and orientation using stratified models, and pairwise interaction terms. Models were adjusted for grade, sexual orientation, race, ethnicity, with both bullying and cyberbullying included as exposures. Persistent sadness and self-harm (binary variables: yes/no) were considered mediators and assessed separately. Prior to mediation analysis, we assessed the strength of associations between suicidality and both self-harming and sadness by performing adjusted multivariable logistic regressions, testing for potential effect modification by sex and sexual orientation via stratified models.

### Mediation analysis

Due to the nonlinear nature of logistic regression, methods for assessing mediated effects either by the product or difference methods do not produce equivalent results(31). As we wish to assess significance of and the mediated proportion of the total effect, we use binomial family probit-link models for the mediation study and first proceed via an adjusted model-based approach for each mediator (sadness and cyberbullying) separately, using the mediation package(32) in the R programming language. In addition, we assess for mediation moderated by sex and by sexual orientation by directly testing the statistical significance of pairwise differences in the Average Causal Mediated Effect (ACME) and Average Direct Effect (ADE) between a chosen level of a covariate and its reference level(32). See Supplemental Appendix for technical details.

## Results

### Participant Demographics

The study population consisted of 4,848 adolescents from Otsego County, New York, who participated in Youth Risk Behavior Surveys (YRBS) between 2016 and 2023 (Table 1). Of these, 51% were female (n=2,476) and 49% were male (n=2,372). Racial composition was predominantly non-Hispanic White (84%). More females identified as bisexual (16.0%) or unsure/questioning (7.3%) than males (4.3% and 4.4%, respectively), who were more likely to identify as heterosexual. Girls reported a higher prevalence of suicidal ideation (14.9% vs. 8.3%), persistent sadness (35.4% vs. 17.2%), and self-harming (23.2% vs. 8.9%) relative to boys. Bullying and cyberbullying were more frequently reported by females (24.2% and 20.8%, respectively) than males (13.6% and 10.8%).

**Table 1.**
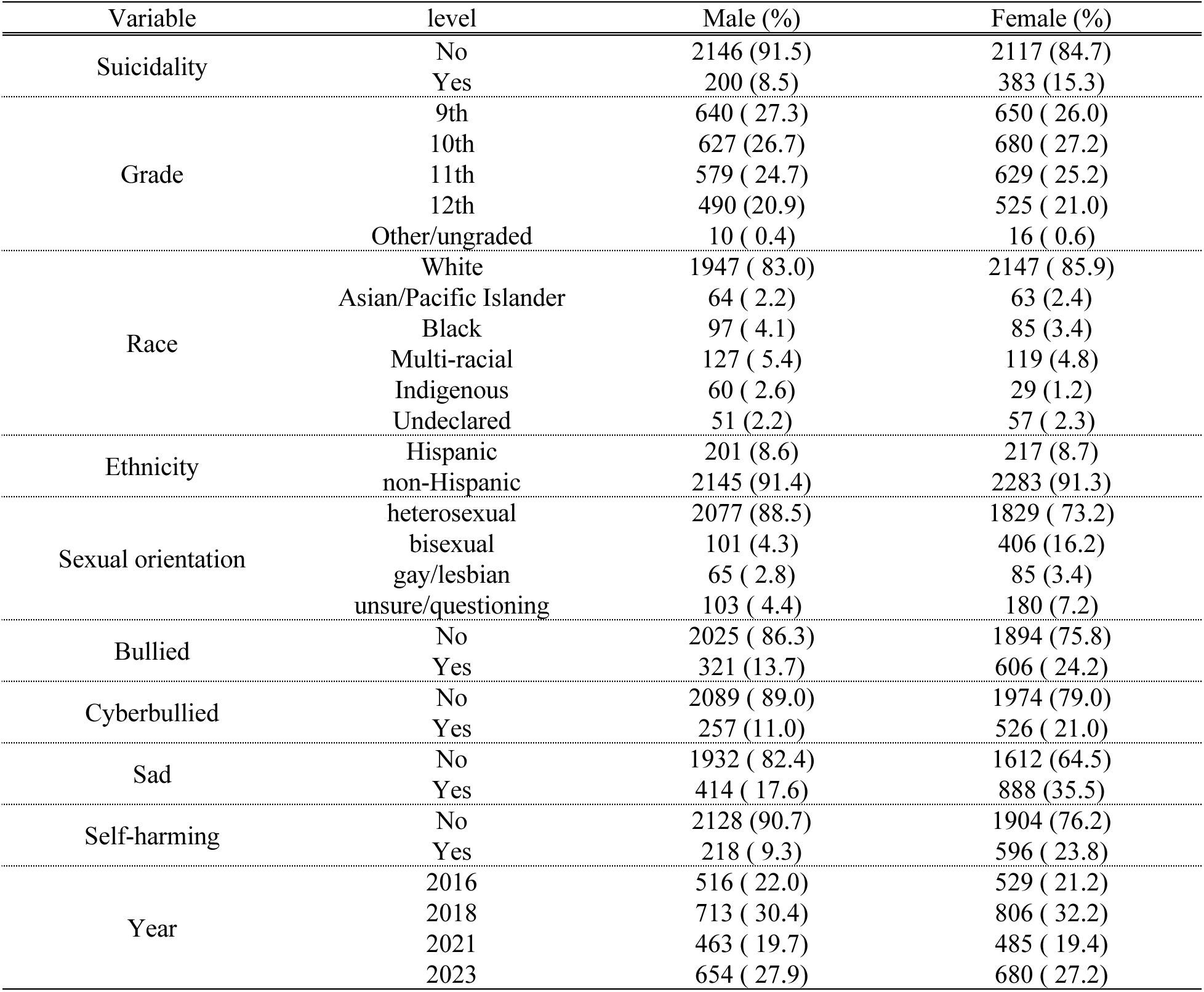
Demographics of study population, 2016-2023. Stratified by sex (females, n=2476; males, n=2,372). Summarized from multiply imputed data, N=4,848. For complete case summary, see Appendix, Table 1.

### Bivariate Analysis

Students experiencing cyberbullying had significantly higher odds of suicidality compared to those who did not experience cyberbullying (OR = 5.0, 95%CI [4.1–6.1], Supplemental Table S2). Similarly, students reporting bullying exposure showed higher odds of suicidality (OR = 4.5, [3.7–5.4]) relative to students who were not bullied. Individuals who reported self-harm exhibited substantially higher odds of suicidality (OR = 20.5, [16.2–26.0]) relative to those who did not self-harm. Females had significantly higher odds of suicidality (OR = 1.9, [1.6–2.3]; Supplemental Table S3), bullying (OR = 2.0, [1.70–2.28]), cyberbullying (OR = 2.17, [1.83–2.53]), and self-harming (OR = 3.1, [2.54–3.54]) and persistent sadness (OR=2.6, [2.3, 3.0]) compared to males. Youth identifying as bisexual (OR = 5.27), gay/lesbian (OR = 4.9), and unsure/questioning (OR = 3.07) had greater odds of suicidality compared to heterosexual youth, with similarly elevated odds for bullying, cyberbullying, persistent sadness and self-harming across all non-heterosexual groups (*p* < 0.001 for all, Table S3). For additional details by race, generally not significant due to a majority white population, see Table S4.

### Multivariable Analysis

In fully adjusted multivariable models, cyber-bullying and conventional bullying were significantly associated with suicidality (OR 2.6, [2.0, 3.2] and OR 2.38 [1.9, 3.0]), as were self-harming (OR 3.3, [2.6, 4.1] and OR 2.1, [1.7, 2.7]) and persistent sadness (OR 3.2, [2.6, 3.9]) and OR 2.0 [1.7, 2.4], for cyberbullying and bullying, respectively). Note that associations with cyberbullying are higher than for conventional bullying (Table 2).

**Table 2.** Pooled associations between suicidality and bullying modalities from logistic regressions run on imputed data (20 imputed datasets). Full models are adjusted for sex, grade, race, study year and sexual orientation. Ethnicity was dropped because it was not significant (91% non-Hispanic population). Exposure reference levels are “No” (not cyber/bullied).

| Outcome | Exposure | Level | OR | 95% CI | p-value |
| --- | --- | --- | --- | --- | --- |
| Suicidality | Cyberbullied | Yes | 2.7 | 2.1, 3.4 | **<0.0001** |
|  | Bullied | Yes | 2.4 | 1.9, 3.0 | **<0.0001** |
| Self-harming | Cyberbullied | Yes | 3.3 | 2.6, 4.1 | **<0.0001** |
|  | Bullied | Yes | 2.1 | 1.7, 2.7 | **<0.0001** |
| Sadness | Cyberbullied | Yes | 3.2 | 2.6, 3.9 | **<0.0001** |
|  | Bullied | Yes | 2.0 | 1.7, 2.4 | **<0.0001** |

### Stratified models

Results from stratified models are consistent by strata across exposures and outcomes as one might expect for potential mediators, with the exception of models stratified by sex, where results for suicidality show higher suicidality estimates among females for cyberbullying than bullying, and the reverse for males (Figure 1, top panels).

**Figure 1.**
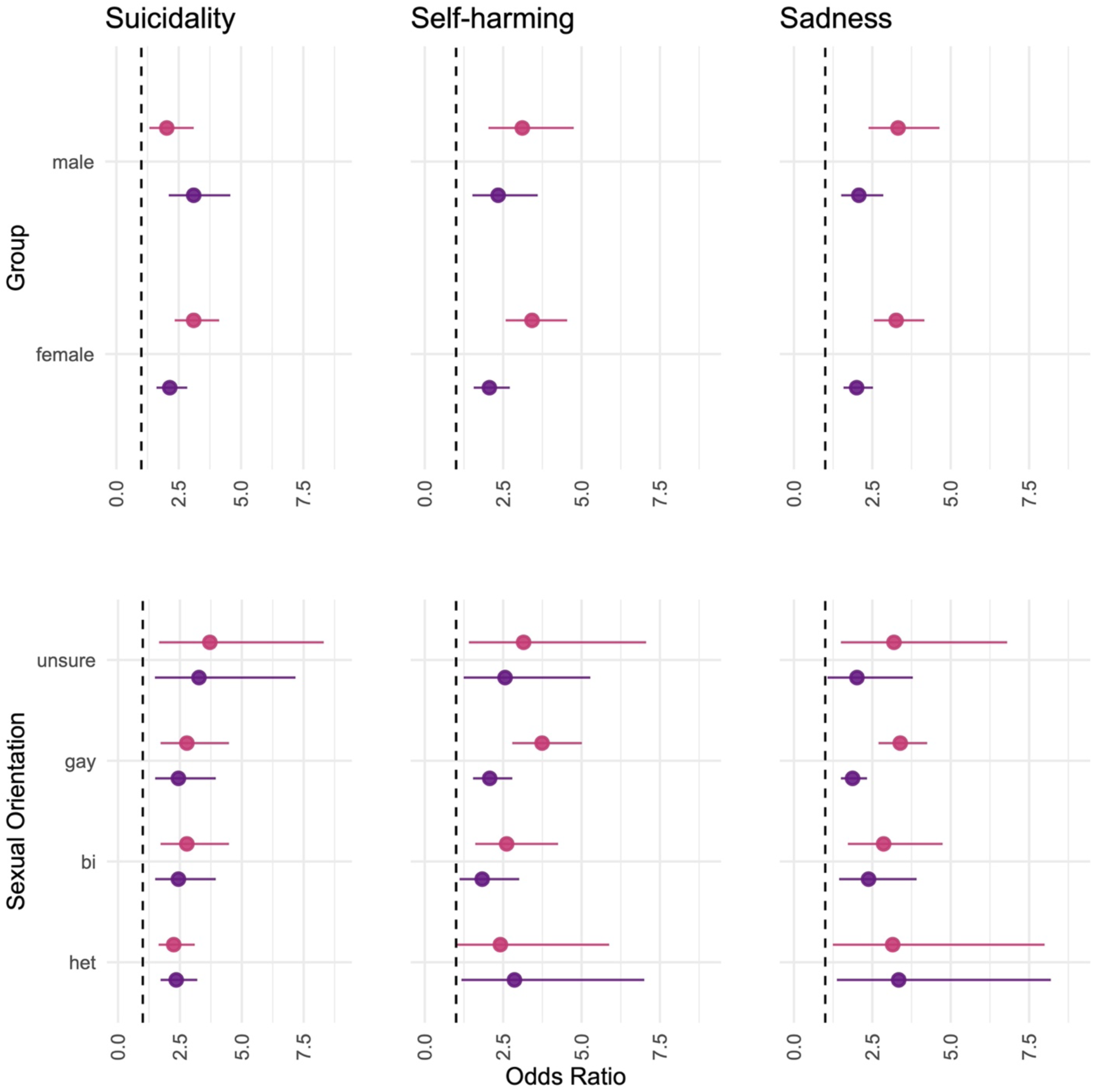
Associations between suicidality, self-harming and sadness with cyberbullying and conventional bullying by sex (top three panels) and by sexual orientation (bottom three panels) from adjusted stratified models. Dashed reference line represents OR=1. Cyberbullying is shown in magenta; bullying in purple.

### Suicidality by sex

Both bullying modalities were strongly associated with suicidality in models stratified by sex. However, we found that associations between suicidality and cyberbullying were stronger in females (aOR: 3.1 [2.3, 4.1] for cyberbullying versus aOR 2.1 [1.6, 2.8] for bullying) than in males, while in males associations between suicidality and conventional bullying were stronger (aOR: 1.9 [1.3, 3.1] for cyberbullying versus OR 2.9 [2.1, 4.6] for bullying; see Table S6 and Figure 1, top panels).

### Self-harming and sadness by sex

We found strong associations between bullying modalities and self-harming in models stratified by sex (Table S6; Figure 1). Odds of self-harming when exposed to either form of bullying were significantly elevated for both males (OR: 3.10 [2.03, 4.74]) and females (OR: 3.32, [2.53, 4.36]) subject to cyberbullying. Notably, females subject to cyberbullying had the highest odds of self-harming, with an OR approaching 3.5. Males also showed increased odds of self-harm in response to cyberbullying, though the estimate was more variable. Similarly, both sexes exhibited increased risk of self-harm when exposed to traditional bullying, with slightly higher estimates in males (i.e., OR: 2.09, [1.57, 2.79] in females; versus OR: 2.33, [1.55, 3.51] in males).

### Suicidality by orientation

In models stratified by orientation, sexual minorities showed slightly higher associations between cyberbullying and suicidality compared to students who identify as heterosexual (Table S6, Figure 1, bottom panels), though confidence intervals overlap due to smaller sample sizes. Students identifying as “unsure/questioning” showed the highest associations between cyberbullying and suicidality (aOR: 4.0, [1.75, 9.1]). Associations between conventional bullying and suicidality had the greatest variability for students who identify as gay/lesbian or “unsure/questioning,” (aOR:2.4, [1.48, 3.93] for gay/lesbian and aOR:3.34, [1.51, 7.3] for unsure/questioning) while conventional bullying was significantly associated with suicidality for heterosexual (aOR: 2.34 [1.7, 3.23]) and bisexual (aOR: 2.41 [1.48, 3.93]) students.

### Self-harming and sadness by orientation

Models stratified by orientation show significant results for all sexual orientations (Table S6; Figure 1). Notably, students identifying as heterosexual showed much higher odds for self-harming (aOR: 3.7; [ 2.69, 5.14]) relative to their odds for suicidality (aOR: 2.3; [1.63, 3.16]) with cyberbullying exposure (Figure 1). Cyberbullied unsure/questioning students also had elevated odds of self-harming (aOR: 3.2 [1.4,7.2]), consistent with their odds for suicidality with exposure. However, under bullying exposure, associations with self-harming are lower (aOR: 2.4, [1.2,4.9]).

### Mediation of suicidality associations by persistent sadness and self-harming

We found substantial and statistically significant mediation by self-harming in associations between suicidality and both cyberbullying and conventional bullying, with higher percentages of mediation for cyberbullying (63% for those experiencing cyberbullying, Figure 2, Table 3) than for bullying (55% for those experiencing bullying). Mediation by persistent sadness was also substantial and significant for both types of bullying, but smaller in proportion than mediation of associations by self-harm. Associations between cyberbullying and suicidality showed 56% mediation by sadness and for conventional bullying, 51% mediation.

**Figure 2.**
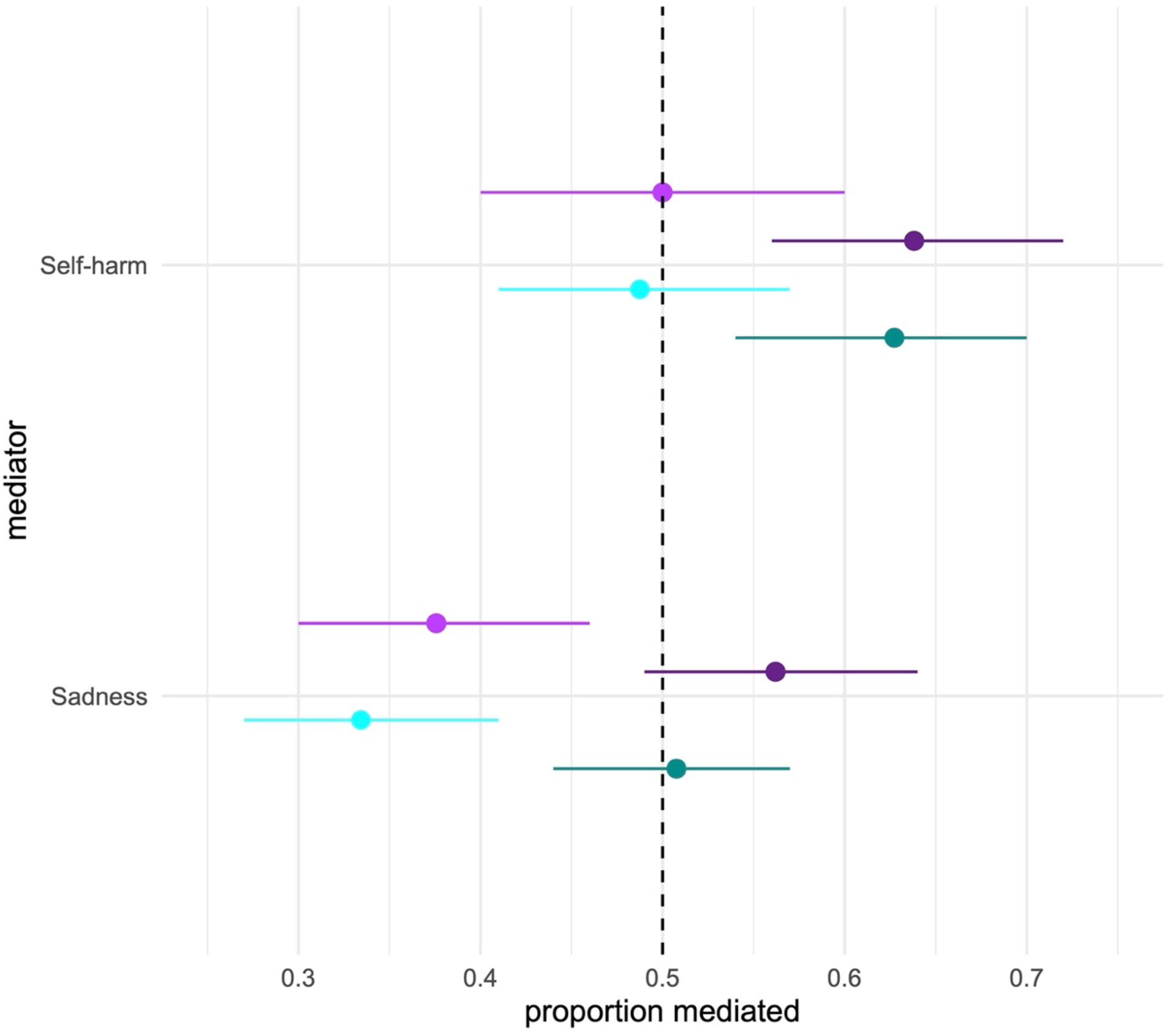
Forest plot visualizing proportion mediation of associations between bullying modalities and suicidality by self-harming and sadness (cyberbullied: dark purple, control: light purple; bullied: dark cyan; control: cyan). Dashed reference line represents a proportion of 0.50 (50%) mediation.

**Table 3.** Results from analysis of associations between bullying modalities and suicidality, mediated by self-harm and persistent sadness respectively, pooled over 20 imputed datasets; shown as proportion mediated (of total effect) with quasi-Bayesian confidence intervals. Models are adjusted for grade, sex, race, sexual orientation, and screen-time (hours), and are binomial (probit link) generalized linear models; (N=4518). Assumes independence of mediators. Please see Supplementary Table S7 for a full listing of mediated (ACME) and direct effects (ADE).

| Mediator | Exposure | Level | proportion mediated (%) | 95% CI | p-value |
| --- | --- | --- | --- | --- | --- |
| Self-harming | Cyberbullying | Not cyberbullied | 0.49 (49) | 0.41, 0.57 | <0.00001 |
|  |  | Cyberbullied | 0.63 (63) | 0.56, 0.70 | <0.00001 |
|  |  | **Average** | **0.58 (58%)** |  |  |
|  | Bullying | Not bullied | 0.41 (41) | 0.37, 0.48 | <0.00001 |
|  |  | Bullied | 0.55 (55) | 0.51, 0.61 | <0.00001 |
|  |  | **Average** | **0.47 (47%)** |  |  |
| Persistent Sadness | Cyberbullying | Not cyberbullied | 0.38 (38) | 0.31, 0.46 | <0.00001 |
|  |  | Cyberbullied | 0.56 (56) | 0.49, 0.64 | <0.00001 |
|  |  | **Average** | **0.47 (47%)** |  |  |
|  | Bullying | Not bullied | 0.33 (33) | 0.27, 0.41 | <0.00001 |
|  |  | Bullied | 0.51 (51) | 0.44, 0.57 | <0.00001 |
|  |  | **Average** | **0.42 (42%)** |  |  |

In addition, assessments of moderated mediation suggest significant positive moderation of the mediation via self-harming by orientation, but not by sex (marginal for cyberbullying, p = 0.08; N/S for bullying, see Supplemental Table S8). For both cyberbullying and bullying, moderation of the average causal mediated effect (ACME) of self-harming was positive and significant for gay, bisexual, and unsure/questioning youth (relative to heterosexual youth, p < 0.0001 for all estimates). Moderation of direct effects in the relationship between cyberbullying and suicidality was also significant for sadness in pairwise comparisons of questioning, bisexual, and gay versus heterosexual youth (p < 0.0001). For bullying, direct effects for both self-harming and sadness were significant in bisexual and gay versus heterosexual youth (p < 0.0001).

## Discussion

Suicidality was significantly associated with bullying modalities, including cyberbullying, and associations were strongly (55% to 63%, depending on bullying type) mediated by both self-harming and persistent sadness in our sample of rural youth. We found that youth who experienced cyberbullying were markedly more likely to experience suicidality than their non-cyberbullied peers, and that the effect differed by sex, a finding consistent with multiple studies demonstrating that cyberbullying is a major psychosocial stressor that exacerbates suicidality risk in adolescents (33, 34). We have discovered significant mediation relationships between bullying and cyberbullying, self-harming, sadness and suicidality in our rurally representative sample.

Our work suggests discrepancies in cyberbullying and bullying victimization and suicidality, particularly among females and sexual minority groups. We found that girls were disproportionately affected by cyberbullying in full and stratified models, with significant mediation of this association by self-harming and sadness. By contrast boys were disproportionately affected by conventional bullying. Still, effect modification of the mediation by sex was marginal (p = 0.08), in contrast with significant effect modification by sex in results from analysis of national YRBS records from a similar period (Odewumi & Jones, 2025). The relatively larger effects of cyberbullying on girls may be due to the prevalence of relational aggression (15), manifesting as gossiping, spreading rumors, friendship betrayals, exclusion, and other manipulative behaviors that are easily facilitated online(35). The increased odds of suicidality among cyberbullied females may also be explained by higher rates of internalizing disorders, such as depression, which is strongly associated with and shown to mediate suicidal behavior (15, 36, 37). By contrast, results found for the national YRBS data (N=94,492) during this period (2015-2023) show stronger associations between suicidality and both bullying modalities in *males* than females from stratified models (Odewumi and Jones, in prep., 2025).

Rural sexual minority youth also showed larger associations between both cyberbullying and bullying and suicidality, though there was no significant effect modification of direct associations by orientation. Prior studies indicate that LGBTQ+ youth are more likely to experience cyberbullying than their heterosexual peers due to discrimination and victimization based on sexual orientation or gender identity (16, 17, 38). Increased exposure to online victimization contributes to higher rates of depressive symptoms, suicidality, and substance abuse (18). The minority stress model (39) suggests that chronic stress due to stigma, discrimination, and victimization leads to adverse mental health outcomes, which might explain heightened suicidality risk among sexual minority youth in our study. In our study of local data, there are differences by sexual orientation, but confidence intervals overlap, and two-way interaction terms between bullying modalities and sexual orientation were not significant. Yet in national data from the same period, sexual orientation is a partial effect modifier in the associations between bullying modalities and suicidality (Odewumi and Jones, in preparation, 2025). In addition, our results from local data for mediation by self-harming show that there is significant effect modification by sexual orientation (p < 0.0001) in the mediation relationships, both in the direct and mediated effects, in pairwise comparisons with the reference level (heterosexual).

Rural youth often face limited access to healthcare, including mental health services, delaying or preventing intervention for those experiencing psychological distress (40). Shortages of mental health providers, long travel distances to healthcare facilities, and financial barriers further restrict access to necessary support (41). Additionally, stigma surrounding mental health issues in rural communities can deter adolescents from seeking help, increasing suicide risk(42). This suggests a critical need for increased mental health resources, school-based educational programs(43), as well as telehealth services in rural regions to support at-risk youth.

Our study suggests that impacts of cyberbullying and bullying on adolescent mental health are complex and intersectional, even in a relatively racially homogeneous population. To effectively address this, schools and communities might implement evidence-based anti-bullying programs to promote digital literacy and peer support (43-47). Expanding access to mental health services, particularly in rural areas, through school-based programs and telehealth initiatives may reduce suicidality among at-risk youth. Additionally, addressing social stigma and fostering inclusive environments for sexual minority youths is essential in mitigating negative effects of cyberbullying and other bullying modalities. Finally, protective factors such as parental and family engagement, formation of supportive peer groups and teaching healthy resilience to youth may mitigate effects of cyber/bullying on adolescent mental health in rural populations.

## Strengths and Limitations

Strengths of this study include that it employs a subset of records from the YRBS surveys performed semi-annually by Bassett Research Institute with local Otsego County school districts from 1995 to 2023, and that the data are multiply imputed to prevent dramatic reduction in sample size. Records are relatively complete even before imputation, giving a snapshot of rural youth sociology and permitting comparisons with a general US population. While it may not be “generalizable” given that the majority of US population is urban and racially diverse, over 60% of US territory is considered rural, and is populated by at-risk farming, logging and fishing communities.

Although this study provides valuable insights into associations between cyberbullying and suicidality in rural populations, it has limitations. The population is racially homogeneous, largely non-Hispanic White. We cannot evaluate the temporality between exposure and outcome because the study uses cross-sectional data. Despite generally high test-retest reliability for YRBS, study variables were self-reported, which may introduce reporting bias. Finally, because YRBS is conducted in schools, it excludes homeschooled or unschooled youth.

## Conclusion

Our study reveals strong associations between bullying modalities, self-harm and suicidality in rural youth from upstate New York, and finds that associations between bullying and cyberbullying victimization and suicidality are significantly mediated by self-harming and sadness. Results suggest disparities in bullying victimization and suicidality, particularly among females and sexual minority groups. Youth experiencing cyberbullying were significantly more likely to experience suicidal ideation, suggesting that cyberbullying is a significant risk factor for suicidality among adolescents. This result is also true for conventional bullying, but cyberbullying was more closely associated with suicidality than conventional bullying in females, while in males, suicidality was more strongly associated with conventional bullying.

These findings emphasize need for targeted, school-based interventions to support vulnerable groups and mitigate mental health risks associated with bullying victimization on and off school campuses.

## Data Availability

Availability of data and material. Records utilized for analysis in this manuscript are maintained and accessed exclusively by the Bassett Research Institute per agreement with the participating school districts. Therefore, sharing of the data beyond our research team is prohibited.

## Acknowledgements

The authors extend our sincere gratitude to the Otsego County school superintendents, school staff, families and students for their participation in the YRBS. We also acknowledge the significant efforts of Aletha Sprague, Bonita Gibb, John May, Anne Gadomski, Julie Dostal and Marion Mossman for their leadership in fostering community partnerships that made YRBS data collection possible. Finally, we thank Melissa Scribani for coordination of YRBS activities, and Megan Kern for assistance with dataset documentation. Critical reviews of the manuscript were provided by Melissa Scribani, Megan Kern, Sarah Otaru, Christiana Odewumi and Max Tweedale.

## Author contributions

LEJ: conceptualization, data curation and cleaning, analytical plan, software, analysis, writing (original draft), manuscript development and revision; RK: writing (original draft); NK: data curation, initial processing, manuscript development. All authors read and approved the final manuscript.

## Availability of data and material

Records utilized for analysis in this manuscript are maintained and accessed exclusively by the Bassett Research Institute per agreement with the participating school districts. Therefore, sharing of the data beyond our research team is prohibited.

## Declarations

### Ethics approval and Consent to Participate

Local YRBS data collection activities and subsequent analysis were approved by the Institutional Review Board of the Mary Imogene Bassett Hospital (Bassett Medical Center) and were conducted in accordance with the ethical standards of the Declaration of Helsinki. Participant consent was provided via voluntary completion of the questionnaire and passive parental permission consent, or opt-out, following the recommended approach by the Centers for Disease Control and Prevention for YRBS.

### Consent for Publication

Not applicable.

## Supplemental Information

**Figure.**
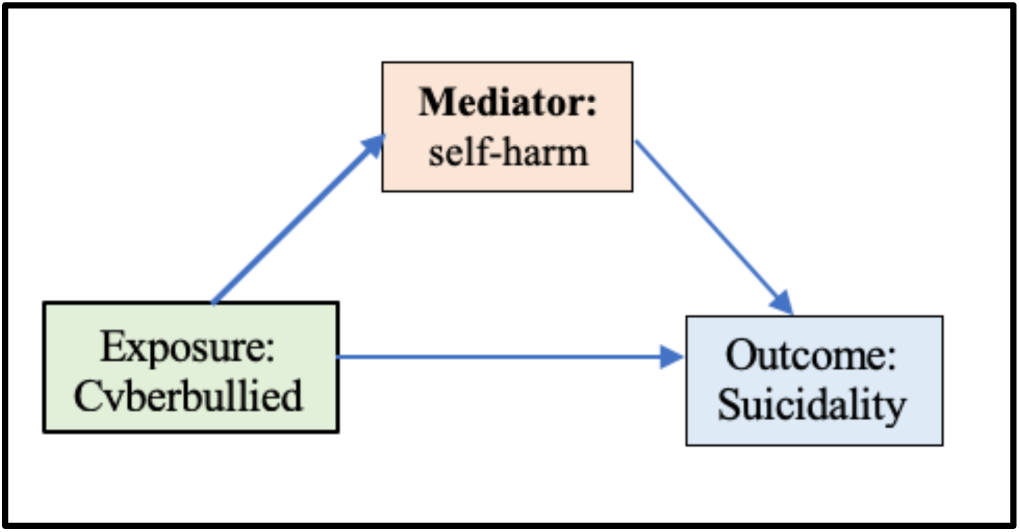
Mediating relationship of self-harming in the association between cyberbullying and suicidality.

**Table S1.** Complete Case Study Population Characteristics (Complete Case, N = 3005). Demographics and characteristics of study population. Counts and proportions of missing responses (NA values) are also reported for each variable.

| Coefficient | level | Female<br>(N=2476) | Male<br>(N=2327) |
| --- | --- | --- | --- |
| cyberbullied (%) | No | 1901 (79.2) | 2003 (89.2) |
|  | Yes | 500 (20.8) | 242 (10.8) |
|  | Missing (NA) | 75 (3.0) | 82 (3.5) |
| Bullied (%) | No | 1829 (75.8) | 1944 (86.4) |
|  | Yes | 585 (24.2) | 306 (13.6) |
|  | Missing | 62 (2.5) | 77 (3.3) |
| Suicidality | No | 2032 (85.1) | 2053 (91.7) |
|  | Yes | 356 (14.9) | 187 (8.3) |
|  | Missing | 88 (3.6) | 87 (3.7) |
| Sex (%) | Female | 2476 (100.0) | - |
|  | Male | - | 2327 (100.0) |
| Grade (%) | 9th | 641 (26.0) | 632 (27.3) |
|  | 10th | 668 (27.1) | 623 (26.9) |
|  | 11th | 625 (25.3) | 573 (24.7) |
|  | 12th | 517 (21.0) | 480 (20.7) |
|  | Other/ungraded | 16 (0.6) | 9 (0.4) |
|  | Missing | 9 (0.4) | 10 (0.4) |
| Race (%) | Asian/Pacific Islander | 63 (2.6) | 61 (2.7) |
|  | Black | 84 (3.4) | 97 (4.2) |
|  | Multi | 116 (4.7) | 127 (5.5) |
|  | Native | 29 (1.2) | 59 (2.5) |
|  | White | 2132 (86.1) | 1935 (83.2) |
|  | Undeclared | 52 (2.1) | 48 (2.1) |
| Ethnicity (%) | Hispanic | 207 (8.5) | 192 (8.5) |
|  | non-Hispanic | 2236 (91.5) | 2075 (91.5) |
|  | Missing | 33 (1.3) | 60 (2.6) |
| Sexual orientation (%) | bisexual | 390 (16.0) | 98 (4.3) |
|  | cis-hetero | 1785 (73.3) | 2033 (88.6) |
|  | gay/lesbian | 83 (3.4) | 62 (2.7) |
|  | unsure/questioning | 178 (7.3) | 102 (4.4) |
|  | Missing | 40 (1.6) | 32 (1.4) |
| Recreational Screen time<br>(hours) | <1 hour | 234 (10.0) | 286 (13.3) |
|  | 1 hour | 238 (10.2) | 230 (10.7) |
|  | 2 hours | 441 (18.9) | 423 (19.7) |
|  | 3 hours | 393 (16.9) | 363 (16.9) |
|  | 4 hours | 279 (12.0) | 203 (9.4) |
|  | 5+ hours | 450 (19.3) | 450 (20.9) |
|  | none | 295 (12.7) | 194 (9.0) |
|  | Missing | 146 (5.9) | 178 (7.6) |

**Table S2.** Crude (unadjusted bivariate) associations between Suicidality (response) and Exposures (cyber bullying, bullying, self-harming). Intercept levels are enclosed in parentheses. Pooling of Chi-square tests results in a D_2_ statistic that is approximately F-distributed (1, 2) and see also (3) for a comparison of pooling methods.

| **Exposure** | **Level** | **OR** | **95% CI** | **p-value** | **$\mathcal{D}_2$, Pr(>$\mathcal{F}$)** |
| --- | --- | --- | --- | --- | --- |
| Bullied | (No) | 0.1 | 0.08, 0.10 | < 0.0001 | $\mathcal{D}_2=271.4$ |
|  | Yes | 4.5 | 3.7, 5.4 | < 0.0001 | p<0.0001 |
| Cyberbullied | (No) | 0.1 | 0.08, 0.10 | < 0.0001 | $\mathcal{D}_2=299.2$ |
|  | Yes | 5.0 | 4.1, 6.1 | < 0.0001 | p<0.0001 |
| Self-harming | (No) | 0.05 | 0.04, 0.07 | < 0.0001 | $\mathcal{D}_2=848.8$ |
|  | Yes | 20.5 | 16.2, 25.9 | < 0.0001 | p<0.0001 |
| Persistent<br>Sadness | (No) | 0.05 | 0.04, 0.07 | < 0.0001 | $\mathcal{D}_2=855.0$ |
|  | Yes | 15.8 | 12.6, 19.6 | < 0.0001 | p<0.0001 |

**Table S3.** Demographic associations. Bivariate associations between sex and sexual orientation and suicidality, cyber bullying, bullying, self-harming, respectively. Intercept levels are enclosed in parentheses. Estimates and Chi-squared statistics pooled on 20 imputed datasets. Pooling of Chi-square tests results in a D_2_ statistic that is approximately F-distributed (1, 2) and see also (3) for a comparison of pooling methods.

| Demographic | Response | Level | OR | 95% CI | p-value | $D_2$,<br>$\Pr(>F)$ |
| --- | --- | --- | --- | --- | --- | --- |
| **Sex** | Suicidality | (male) | 0.09 | 0.08, 0.11 | <0.0001 | $\mathcal{D}_2=47.41$ |
|  |  | Female | 1.9 | 1.6, 2.3 | <0.0001 | p <0.0001 |
| | Bullied | (male) | 0.16 | 0.14, 0.18 | <0.0001 | $\mathcal{D}_2=84.02$ |
|  |  | Female | 2.03 | 1.70, 2.28 | <0.0001 | p <0.0001 |
| | Cyberbullied | (male) | 0.12 | 0.11, 0.14 | <0.0001 | $\mathcal{D}_2=86.38$ |
|  |  | Female | 2.17 | 1.83, 2.53 | <0.0001 | p <0.0001 |
| | Self-Harming | (male) | 0.11 | 0.09, 0.12 | <0.0001 | $\mathcal{D}_2=142.85$ |
|  |  | Female | 3.1 | 2.54, 3.54 | <0.0001 | p <0.0001 |
| | Persistent sadness | (male) | 0.21 | 0.19, 0.24 | <0.0001 | $\mathcal{D}_2=192.72$ |
|  |  | Female | 2.6 | 2.3, 3.0 | <0.0001 | p <0.0001 |
| **Sexual Orientation** | Suicidality | (heterosexual) | 0.09 | 0.08, 0.10 | <0.0001 |  |
| | | bisexual | 5.27 | 4.21, 6.59 | <0.0001 | $\mathcal{D}_2=94.5$ |
|  |  | gay/lesbian | 4.88 | 3.35, 7.00 | <0.0001 | p <0.0001 |
|  |  | unsure/questioning | 3.07 | 2.25, 4.13 | <0.0001 |  |
|  | Bullied | (heterosexual) | 0.19 | 0.18, 0.21 | <0.0001 |  |
| | | bisexual | 2.14 | 1.72, 2.65 | <0.0001 | $\mathcal{D}_2=37.0$ |
|  |  | gay/lesbian | 3.29 | 2.32, 4.66 | <0.0001 | p <0.0001 |
|  |  | unsure/questioning | 2.29 | 1.76, 3.0 | <0.0001 |  |
|  | Cyberbullied | (heterosexual) | 0.16 | 0.14, 0.17 | <0.0001 |  |
| | | bisexual | 2.57 | 2.07, 3.19 | <0.0001 | $\mathcal{D}_2=34.5$ |
|  |  | gay/lesbian | 3.16 | 2.17, 4.61 | <0.0001 | p <0.0001 |
|  |  | unsure/questioning | 1.52 | 1.11, 2.08 | 0.01 |  |
|  | Self-harming | (heterosexual) | 0.13 | 0.12, 0.14 | <0.0001 |  |
| | | bisexual | 6.4 | 5.1, 8.1 | <0.0001 | $\mathcal{D}_2=100.5$ |
|  |  | gay/lesbian | 5.18 | 3.55, 7.56 | <0.0001 | p <0.0001 |
|  |  | unsure/questioning | 2.87 | 2.05, 4.0 | <0.0001 |  |
|  | Persistent sadness | (heterosexual) | 0.26 | 0.24, 0.29 | <0.0001 |  |
| | | bisexual | 5.0 | 4.1, 6.1 | <0.0001 | $\mathcal{D}_2=122.3$ |
|  |  | gay/lesbian | 4.5 | 3.2, 6.3 | <0.0001 | p <0.0001 |
|  |  | unsure/questioning | 2.54 | 1.97, 3.3 | <0.0001 |  |

**Table S4.** Demographic associations. Bivariate associations between Race and cyber bullying, bullying, and self-harming, respectively. Intercept terms are enclosed in parentheses. Estimates and Chi-squared statistics pooled on 20 imputed datasets. Pooling of Chi-square tests results in a D_2_ statistic that is approximately F-distributed (1).

| Response | Predictor: Race | OR | 95% CI | p-value | $D_2$,<br>$\Pr(>\mathcal{F})$ |
| --- | --- | --- | --- | --- | --- |
| Suicidality | (Non-Hispanic White) | 0.12 | 0.11, 0.13 | <0.0001 | $\mathcal{D}_2=4.54$<br>$p < 0.001$ |
|  | Asian/Pacific Islander | 1.52 | 0.86, 2.48 | 0.11 |  |
|  | Black | **1.62** | **1.1, 2.45** | **0.025** |  |
|  | Multiracial | **1.87** | **1.3, 2.6** | **0.0003** |  |
|  | Native | **1.91** | **1.1, 3.4** | **0.026** |  |
|  | Undeclared | 1.42 | 0.80, 2.55 | 0.23 |  |
| Bullied | (Non-Hispanic White) | 0.23 | 0.21, 0.25 | <0.0001 | $\mathcal{D}_2=2.34$<br>0.04 |
|  | Asian/Pacific Islander | 0.7 | 0.5, 1.3 | 0.37 |  |
|  | Black | 1.00 | 0.66, 1.7 | 0.95 |  |
|  | **Multiracial** | **1.41** | **1.04, 1.92** | **0.03** |  |
|  | **Native** | **1.84** | **1.13, 3.0** | **0.01** |  |
|  | Undeclared | 1.04 | 0.63, 1.7 | 0.88 |  |
| Cyberbullied | (Non-Hispanic White) | 0.18 | 0.17, 0.20 | <0.0001 | $\mathcal{D}_2=2.72$<br>0.02 |
|  | Asian/Pacific Islander | 1.1 | 0.66, 1.74 | 0.79 |  |
|  | Black | 1.42 | 0.96, 2.1 | 0.08 |  |
|  | **Multiracial** | **1.52** | **1.1, 2.1** | **0.01** |  |
|  | **Native** | **1.74** | **1.04, 2.9** | **0.03** |  |
|  | Undeclared | 1.12 | 0.66, 1.9 | 0.66 |  |
| Self-harming | (Non-Hispanic White) | 0.20 | 0.18, 0.21 | <0.0001 | $\mathcal{D}_2=.77$<br>0.57 |
|  | Asian/Pacific Islander | 0.94 | 0.54, 1.63 | 0.83 |  |
|  | Black | 1.14 | 0.75, 1.63 | 0.55 |  |
|  | Multiracial | 1.14 | 0.79, 1.63 | 0.48 |  |
|  | Native | 1.47 | 0.85, 2.53 | 0.17 |  |
|  | Undeclared | 1.35 | 0.81, 2.24 | 0.12 |  |
| Persistent sadness | (Non-Hispanic White) | 0.35 | 0.33, 0.38 | <0.0001 | $\mathcal{D}_2=1.95$<br>0.08 |
|  | Asian/Pacific Islander | 1.1 | 0.75, 1.68 | 0.57 |  |
|  | Black | 1.3 | 0.94, 1.81 | 0.12 |  |
|  | **Multiracial** | **1.4** | **1.1, 1.9** | **0.01** |  |
|  | Native | 1.1 | 0.68, 1.8 | 0.7 |  |
|  | Undeclared | 1.2 | 0.8, 1.9 | 0.3 |  |

**Table S5.**
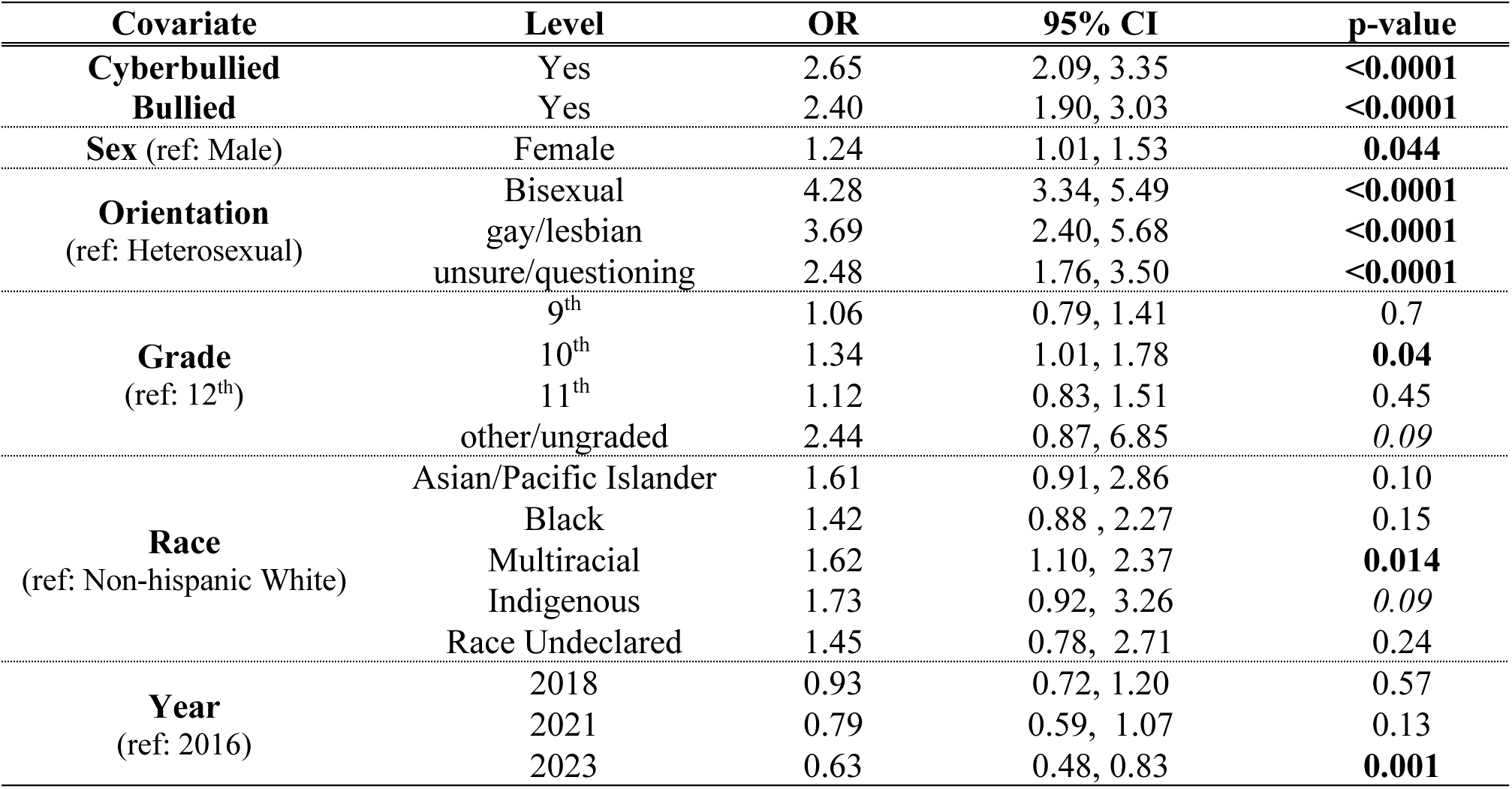
Pooled associations between self-harming and bullying modalities from logistic regressions run on imputed data (20 imputed datasets). Models are adjusted for sex, grade, race, and sexual orientation. Ethnicity was dropped because it was not significant.

**Table S6.**
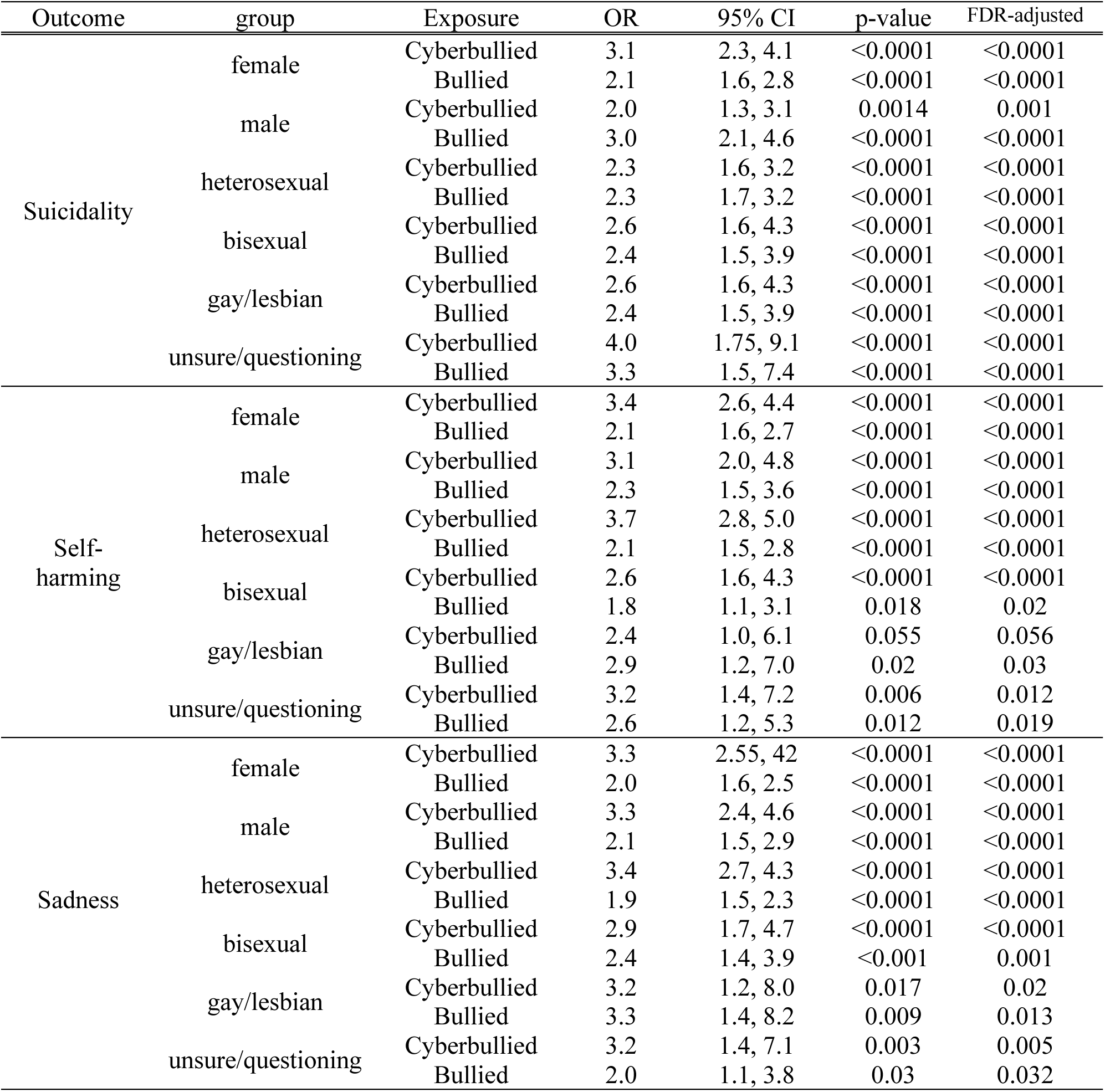
Associations between bullying modalities (cyberbullying, traditional in-person school bullying) and odds of suicidality and self-harming from adjusted logistic regression models stratified on sex or sexual orientation; results pooled over 20 imputed datasets. Note that interaction terms between sex or orientation and bullying modalities were not significant in full adjusted models.

**Table S7.**
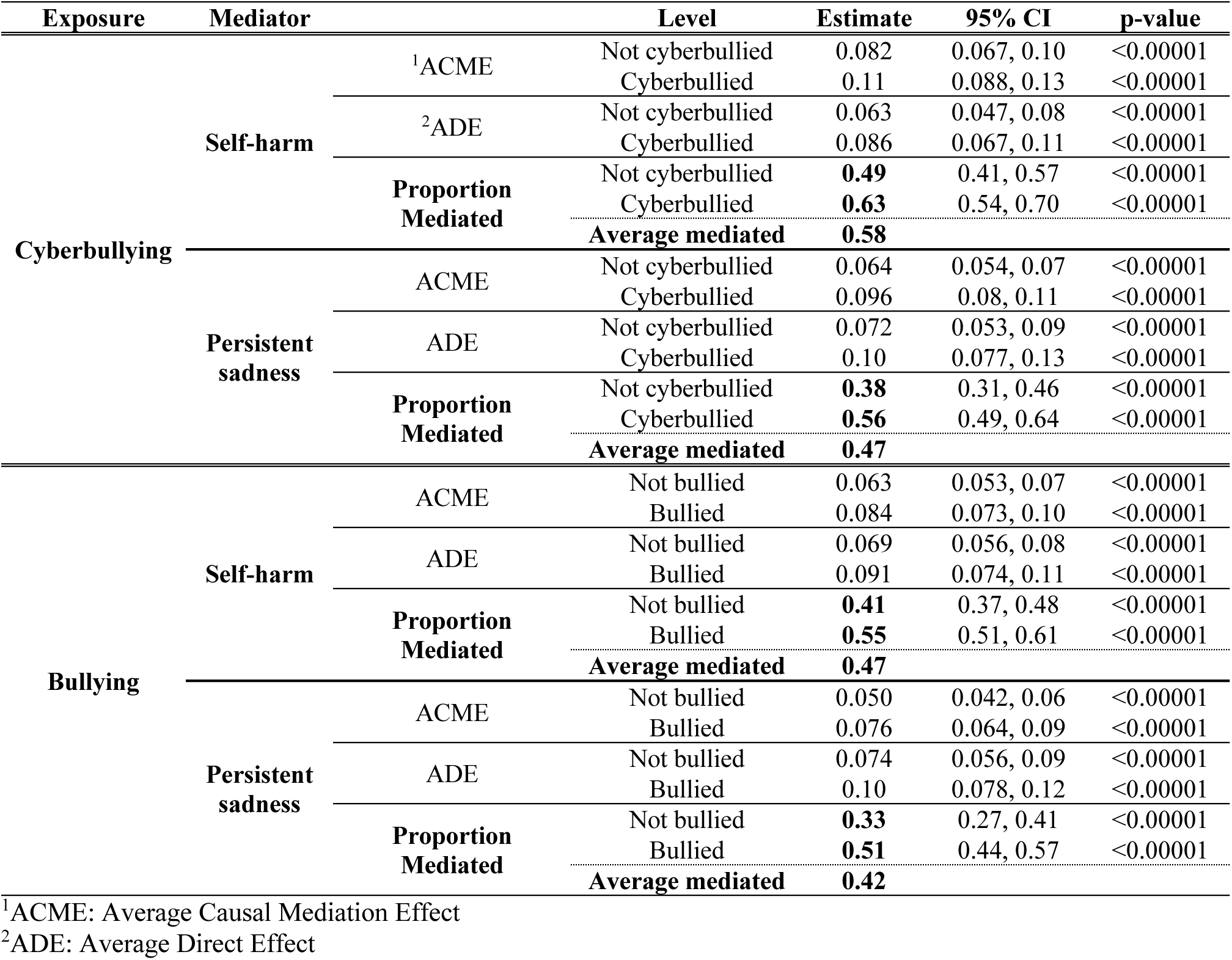
Mediation Analysis. Results from mediation analysis of adjusted associations between bullying modalities and suicidality, mediated by self-harming or persistent sadness (sadness), pooled over 20 imputed datasets shown with quasi-Bayesian confidence intervals. Assumes independence between bullying modalities. Models are adjusted for grade, sex, race, sexual orientation, and screen-time (hours), and are binomial (probit link) generalized linear models (N=4518). The figure box below is a simple example rendering of the mediation relationship.

**Table S8.**
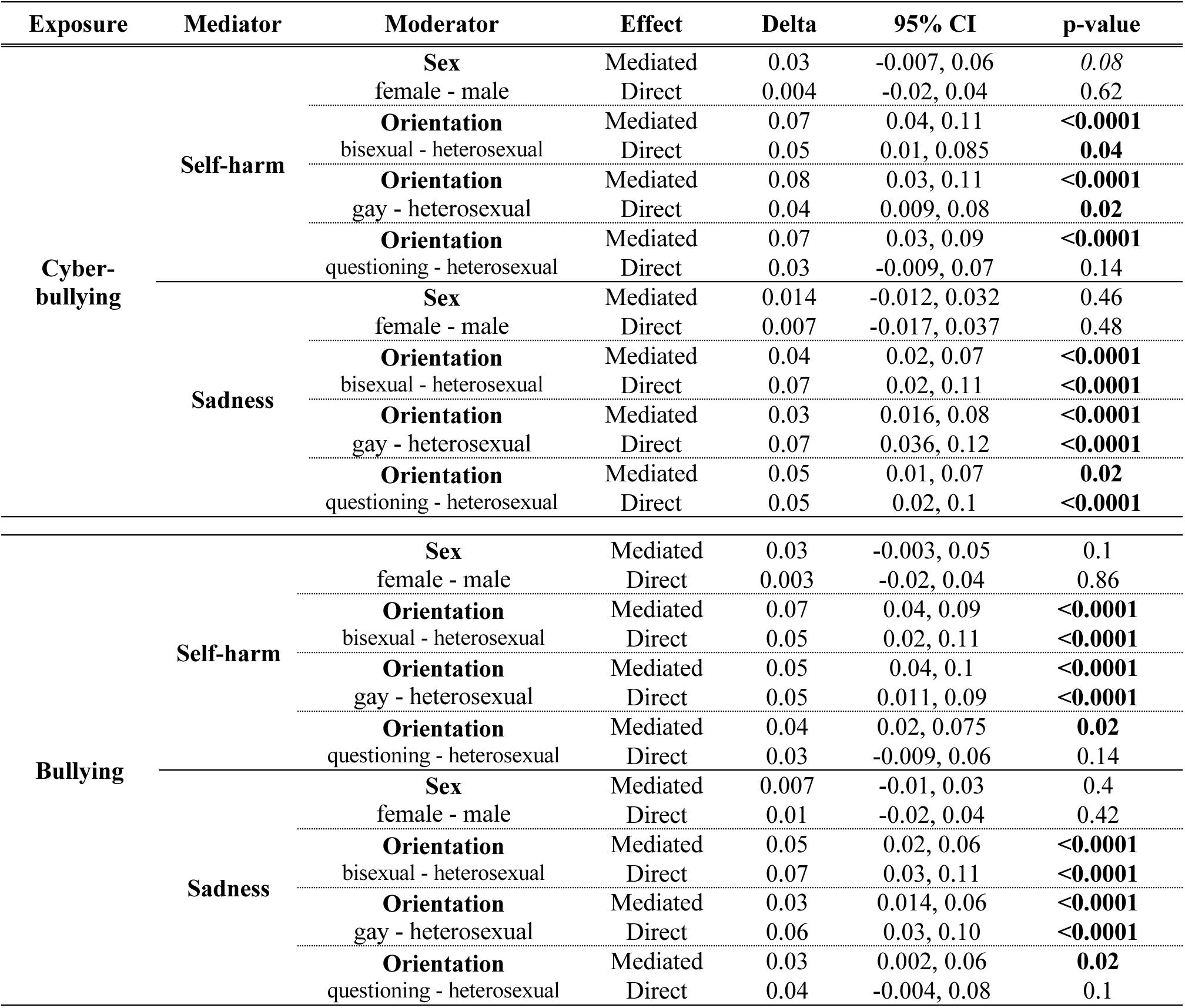
Moderated mediation analysis. Results from moderated mediation analysis of associations between bullying modalities and suicidality, mediated by self-harm, pooled over 20 imputed datasets shown with quasi-Bayesian confidence intervals. We tested for effect modification of both mediated and direct effects by sex and sexual orientation. Effect modification is assessed pairwise by differencing estimates for the reference level (e.g., heterosexual) from estimates for the level of interest (e.g., gay) for Mediated (ACME) and Direct (ADE) effects; this difference is labeled ‘Delta’ in the table below. Note that the link function is probit, not logit, so results should not be compared with OR estimates.

## Appendix 1

### Mediation analysis

Because of the nonlinear nature of logistic regression, methods for calculating mediated effects either by the product or difference methods do not produce equivalent results (4). Since our interest is in establishing existence of and assessing proportion of mediation rather than directly comparing with results from our logistic models, we use binomial family probit-link models for the mediation study. Probit-link binomial family models are often used in lieu of logit-link models in mediation due to better coefficient standardization and easier interpretation of mediated effects. We proceed via a model-based approach, first creating two models, one to assess adjusted associations between the mediator (self-harming) and exposure (bullying modalities), and a second to model the conditional distribution of the outcome (suicidality) given bullying modalities, the mediator, and covariates (5, 6). The fitted models are fed to the ‘mediate’ function, which calculates estimates of the Average Causal Mediation Effect (ACME), the Average Direct Effect (ADE) and other quantities via algorithms described in Imai et al. (6, 7). We address the bullying modalities as separate exposures here, using a quasi-bayesian method with 1000 bootstrap simulations per imputed dataset. Standard errors are calculated via a robust sandwich method using White’s heteroskedasticity-consistent estimator for the covariance matrix from the sandwich package (8).

All 20 datasets are passed through this workflow and the results pooled. In addition, we assess for mediation moderated by sex and by sexual orientation by directly testing the statistical significance of pairwise differences in the Average Causal Mediated Effect (ACME) and Average Direct Effect (ADE) between a chosen level of a covariate and its reference level (5). For example, to assess moderation by sex of mediation via self-harming, we difference the ACME we find for females from that of males, then test for statistical significance as described in Tingley et al. (5).

